# Reliability and accuracy of medication-based performance measures for assessment of outpatient cardiology clinicians

**DOI:** 10.64898/2026.09.29.26364366

**Authors:** Christine Chow, Elise C. Gunzburger, Chris Wilson, Taufiq S. Salahuddin, Paul L. Hess, Utibe R. Essien, Fatima Rodriguez, Kai-Chun Yang, Boback Ziaeian, Jacob A. Doll

## Abstract

**Background:** Multiple performance measures are used to assess the management of common cardiovascular conditions, such as coronary artery disease (CAD), heart failure with reduced ejection fraction (HFrEF), and atrial fibrillation (AF). These measures are sometimes used to assess individual clinicians, despite lack of evidence of reliability and accuracy. We used national Veteran Affairs (VA) data to test medication-based performance measures for assessment of clinician performance in outpatient cardiology care.

**Methods:** This retrospective study examined VA cardiology clinician/Veteran dyads from 2017- 2023 to evaluate 10 medication-based performance measures for CAD, HFrEF, or AF with elevated stroke risk. Achievement of each performance measure was defined as an active prescription of an indicated medication for each eligible patient encounter, summarized at the clinician level. Reliability was calculated using the beta-binomial method. A Bayesian causal model was used to assess the impact of case complexity on performance measure achievement. Finally, we performed peer review of a subset of cases (n = 952) to assess the accuracy of three representative performance measures.

**Results:** Of 456,071 unique cardiology encounters included in this study, 312,331 included CAD, 224,599 included HFrEF, and 178,678 included AF. These patients were treated by 1,043 cardiology clinicians. Median performance measure achievement ranged from 0.23 (SGLT2i for HFrEF) to 0.99 (beta blocker for HFrEF). Measure reliability varied from 0.63 to 0.90. Case complexity did not significantly affect performance measure achievement. Using peer review as the gold standard, accuracy was poor to moderate (balanced accuracy 0.51-0.73), and measures underestimated performance relative to peer review.

**Conclusions:** Ten medication-based performance measures for CAD, HFrEF, and AF can be efficiently measured using electronic health record data, and most have sufficient reliability to support clinician benchmarking and the identification of potential care gaps without need for risk adjustment. However, suboptimal accuracy compared with a peer review gold standard indicates that these measures may be insufficient as a standalone assessment of clinician quality.

## Introduction

Multiple performance measures are used to assess the management of common cardiovascular conditions such as coronary artery disease (CAD), heart failure with reduced ejection fraction (HFrEF), and atrial fibrillation (AF)^1–3^. There are potential advantages to applying these measures to individual clinician performance. Clinician-level performance data can be used to identify educational and quality improvement opportunities^4–6^. Performance measures can meet regulatory requirements to monitor clinician competency^7,8^. Audit and feedback has a positive impact on performance^9^. Many health systems already use performance measures for clinician assessment. For example, the Center for Medicare and Medicaid Services (CMS) Merit-Based Incentive Program (MIPS) includes clinician-level performance indicators that impact reimbursement, and the Veterans Affairs (VA) Health Care System mandates the use of performance measures for the ongoing, routine assessment of cardiology clinicians^10^.

However, there are several limitations to applying cardiovascular performance measures to individuals: sample sizes are smaller compared with measurement of hospital performance; current measures often require chart abstraction and/or peer review which can be labor-intensive; measures that are commonly applied to hospitals have not, in most cases, been tested for reliability at the clinician level^11,12^. The distribution of performance and appropriate thresholds for outlier identification are unknown, especially for clinicians with low case volume. Finally, the impact of case mix on clinician measure achievement has not been established^13,14^. Most clinician-level performance measures in current use are not risk adjusted, but there are concerns that clinicians who care for high-risk and complex patient panels will be penalized by unadjusted performance measures.

We therefore sought to determine if cardiovascular measures evaluating the use of guideline-directed medical therapy could be assessed using electronic health record (EHR) and pharmacy data, whether these measures are accurate and reliable at the individual clinician level, and if risk adjustment for patient factors significantly modified achievement. To achieve this goal, we used a national cohort of Veterans with cardiovascular disease and VA clinicians.

## Methods

### Population

We evaluated a cohort of cardiology clinicians and patients with cardiovascular disease from 2017-2023 in the VA Health Care System. To assign performance measures to individual clinicians, we first identified clinician/patient dyads with a longitudinal treatment relationship. We identified patients with cardiac conditions that are currently assessed by performance measures, including: 1) HFrEF, identified by ICD9/10 codes for systolic heart failure and then limited to patients with most recent left ventricular ejection fraction (LVEF) ≤40%^15,16^; 2) CAD, identified by ICD9/10 codes^17^, and 3) atrial fibrillation or flutter with elevated risk of stroke defined as a CHA2DS2-VASc score of ≥2 for men and ≥3 for women. Cardiology outpatient visits with these patients were identified using administrative stop codes, and the primary clinician for each visit was identified. Of these clinicians, we excluded any VA employee who was not identified as a physician, nurse practitioner (NP), or physician assistant (PA) using VA staff tables. We then identified longitudinal relationships between patients and individual clinicians as defined by ≥2 encounters within 1 year or ≥3 encounters within 3 years. Finally, to ensure all clinicians had adequate case volume for performance assessment, we limited our clinician cohort to individuals with at least 50 annual eligible outpatient encounters for the period they were active in VA^12^. The patient cohort was then defined as all patients with eligible cardiology encounters with these clinicians. If a patient met eligibility for a clinician-patient relationship with more than one clinician (which could include either sequential or overlapping patterns of care), performance measure achievement for each eligible encounter was assigned to the primary clinician for that encounter.

### Data definitions

Hospital, clinician, and patient data were obtained from VA Corporate Data Warehouse (CDW) and the Consolidated Data Set^18^. This included pharmacy, laboratory, medication allergy/intolerance, and LVEF data to evaluate eligibility for individual performance measures. Cardiovascular conditions and medical co-morbidities were defined by administrative diagnostic and procedure codes, with at least 1 inpatient or 2 outpatient codes present during or prior to the encounter of interest. Contraindications due to drug allergies or intolerances were applied for any applicable allergy present prior to or during the encounter of interest; allergies or intolerances to one medication in a class were applied to all medications in that class, except for statins. Statins were considered contraindicated if two or more statins were listed on the allergy/intolerance list. Renal function contraindications were applied for the most recent applicable laboratory value prior to or on the day of the encounter of interest. LVEF was assessed using a natural language processing algorithm developed and tested with VA data^19^.

The encounter ejection fraction LVEF was the most recent LVEF captured from an echocardiogram or nuclear study within 2 years of the encounter.

Self-reported patient race/ethnicity was categorized into White, Asian, American Indian or Alaska Native, Black or African American, Hispanic or Latino, Native Hawaiian or Other Pacific Islander, and those who listed more than one race, per VA guidance at the time of the analysis. Clinician characteristics were obtained from CDW staff tables.

### Performance measures

Candidate performance measures were selected from mandatory Ongoing Professional Practice Evaluation (OPPE) metrics from VA, measures endorsed by cardiology clinical societies, and emerging measures that included the use of novel medication classes, such as sodium glucose cotransporter 2 (SGLT2) inhibitors, glucagon-like peptide 1 (GLP1) agonists, and angiotensin receptor-neprilysin inhibitors (ARNI) that were newly recommended in society guidelines but not yet included in formal measure sets^1–3^. For each candidate measure, we determined the feasibility of assessment using existing VA EHR data, including ability to define an appropriate population, exclude patients with contraindications, and assess outcomes using medication prescription data. Ten measures were selected for additional testing (**Table 1**).

**Table 1.** Median clinician level achievement and reliability of outpatient cardiology performance measures for patients with atrial fibrillation, coronary artery disease, and heart failure with reduced ejection fraction.

| Condition | Measures | Median Performance | Reliability |
| --- | --- | --- | --- |
| AF | Anticoagulation for patients with elevated stroke risk | 0.82 | 0.63 |
| CAD | Any statin therapy | 0.89 | 0.64 |
| CAD | Any ACEI/ARB/ARNI | 0.73 | 0.66 |
| CAD | Any Beta blocker | 0.89 | 0.73 |
| CAD | SGLT2i or GLP1 for patients with diabetes | 0.32 | 0.90 |
| HFrEF | Any ACEI/ARB/ARNI | 0.75 | 0.63 |
| HFrEF | ARNI | 0.26 | 0.86 |
| HFrEF | Any Beta Blocker | 0.99 | 0.75 |
| HFrEF | Any MRA | 0.45 | 0.74 |
| HFrEF | Any SGLT2i | 0.23 | 0.90 |
| **Composite Measures** |  |  |  |
| Overall composite (10 measures) |  | 0.72 | 0.60 |
| CAD Composite (4 measures) |  | 0.75 | 0.76 |
| HFrEF Composite (5 measures) |  | 0.61 | 0.59 |

**Abbreviations:**
AF: atrial fibrillation or flutter with increased stroke risk
CAD: coronary artery disease
HFrEF: heart failure with reduced ejection fraction
ACEI: angiotensin-converting enzyme inhibitor
ARB: angiotensin II receptor blockers
ARNI: angiotensin receptor-neprilysin inhibitor
SGLT2i: sodium-glucose cotransporter-2 inhibitor
GLP1: glucagon-like peptide-1 receptor agonist
MRA: mineralocorticoid receptor antagonist

Each performance measure was calculated independently for each eligible clinical encounter and assigned to the primary clinician for that encounter. An eligible encounter was defined as any cardiology visit from 2017-2023 with a patient/clinician dyad defined above and including at least one of the three cardiology conditions of interest (CAD, HFrEF, AF). Patients with multiple eligible encounters with an individual clinician were limited to one encounter per year, chosen at random, to avoid biasing the measure by repeated assessments of high-volume care users.

Clinician-level performance achievement was calculated using an opportunity-based approach; performance achievement was calculated as the rate of all encounters where the measure was achieved divided by all eligible encounters. Clinicians were therefore assigned a separate performance achievement rate for each of the 10 measures, with different numerators/denominators for each measure determined by the distribution of eligible conditions in their patient cohort.

### Statistical analysis

Medians and interquartile ranges were calculated for each performance measure, and distributions were represented as box plots. We assessed the reliability of each performance measure separately using the beta-binomial methodology endorsed by the National Quality Forum for the testing of clinician-level performance measures^20^. Reliability represents the consistency and reproducibility of measurements. For each measure, we assigned each eligible encounter a pass/fail value with clinician-level performance summarized as a rate between 0 and 1. We then ran a beta-binomial model to assess clinician-to-clinician variance and clinician-specific variance. These values were used to calculate a clinician-level reliability value. The overall reliability for each measure was calculated as the mean reliability for all included clinicians.

We sought to determine if adjustment for case complexity would significantly impact the distribution of clinician performance in our sample. For representation of patient complexity, we used the VA Care Assessment Needs (CAN) score, a well-validated predictor of mortality and hospital admission risk for VA patients^21^. To estimate the potential impact of case mix on clinician success rate, we fit a series of Bayesian causal models with the goal of assessing the strength and directionality of patient complexity on each of the proposed measures. Once the model was fit on observed patients, we simulated an intervention by increasing patient complexity (CAN score) by a standard deviation. We then sampled from the posterior predictive distribution with both actual patients and patients with artificially increased complexity and measured the difference of impact on outcomes, holding all other coefficients fixed. We then repeated all models using Elixhauser comorbidity index as an alternative measure of complexity.

We then sought to determine if a composite measure could be developed to summarize care within clinical conditions or across all 10 measures. We first assessed correlations between performance measures using Pearson correlation coefficients analyzed at the clinician level, summarized from 0-1 with a value of 1.0 representing perfect correlation. A value of 0.5 or greater is considered a moderate to strong correlation. We then attempted to construct a weighted composite measure, but use of principal components analysis and factor analysis did not identify underlying patterns in the data supportive of consolidating or weighting individual measures^22,23^. We therefore elected to develop an unweighted opportunity composite, measured at the clinician level, which was defined as the total times all eligible performance opportunities were met divided by the total number of opportunities (with each assessed encounter contributing between 1 and 10 opportunities depending on the number of eligible measures). Similar composite measures were developed for the CAD and HFrEF measures. The reliability of these composite measures was tested using the same methods described above.

Finally, we selected a sample of encounters to compare the electronic heath record derived medication performance measures to case-based peer review performed by VA cardiology clinicians. 57 clinicians performed 952 reviews, of which 423 included AF, 647 included CAD, and 521 included HFrEF. Peer reviewers accessed de-identified case documentation and related clinical data (allergies, vital signs, laboratories values, medication lists) using a secure website and submitted assessments of performance measure achievement within the website. Three performance measures were selected for peer review: 1) Anticoagulation for patients with AF and elevated stroke risk; 2) Statin therapy for patients with CAD; 3) Angiotensin-converting enzyme inhibitors (ACEI)/angiotensin II receptor blockers (ARB)/ARNI for patients with HFrEF. A 2x2 contingency table was constructed for each measure, from which we calculated: 1) sensitivity and specificity relative to peer review; 2) positive and negative predictive value; 3) balanced accuracy, defined as the mean of sensitivity and specificity (range 0-1, with 0.5 indicating no predictive value); 4) Cohen’s kappa (0.01-0.20 slight agreement, 0.21-0.40 fair agreement, 0.41-0.60 moderate agreement, 0.61-0.80 substantial agreement, and 0.81-1.00 almost perfect agreement). We also assessed for any systematic under- or overestimation of performance relative to peer review with a directional bias test (false positives – false negatives). Some encounters were assessed by two peer reviewers, so observations were not fully independent and confidence intervals were therefore generated using a cluster bootstrap including 2,000 resamples drawn at the encounter level with replacement.

## Results

Of 456,071 unique cardiology encounters included in this study, 312,331 included CAD, 224,599 included HFrEF, and 178,678 included AF with elevated stroke risk. These patients were treated by 1,046 cardiology clinicians. Among all clinicians, 51.0% were women (**Table 2**), and 53.1% were physicians, 9.3% PAs, and 37.6% NPs. The total number of assessed encounters ranged from 52-11,568 (median 758) across all 1,046 clinicians.

**Table 2.** Clinician characteristics.

| **Characteristic** | **All**<br>**1043 (100%)** |
| --- | --- |
| Clinician Sex |  |
| Male | 490 (47.0%) |
| Female | 532 (51.0%) |
| Unknown | 21(2.0%) |
| Clinician Training |  |
| Physician | 554 (53.1%) |
| Nurse Practitioner | 392 (37.6%) |
| Physician Assistant | 97 (9.3%) |
| Encounters |  |
| AF | 178678 (39.2%) |
| CAD | 312331 (68.5%) |
| HFrEF | 224599 (49.2%) |
| Patient volume,<br>median [IQR] |  |
| AF | 54 [19, 140] |
| CAD | 81 [29, 209] |
| HFrEF | 69 [25, 180] |

Abbreviations:
AF: atrial fibrillation or flutter with increased stroke risk
CAD: coronary artery disease
HFrEF: heart failure with reduced ejection fraction
IQR: interquartile range

Performance measure achievement varied by measure (**Table 1**), ranging from a median of 23.1% (SGLT2i for HFrEF) to 98.6% (beta blocker for HFrEF). At the clinician level, there was significant variation in performance measure achievement, most notable among the emerging performance measures including ARNI, SGLT2i, and GLP1a, in addition to the mineralocorticoid receptor antagonist (MRA) for HFrEF measure (**Figure 1**).

**Figure 1.**
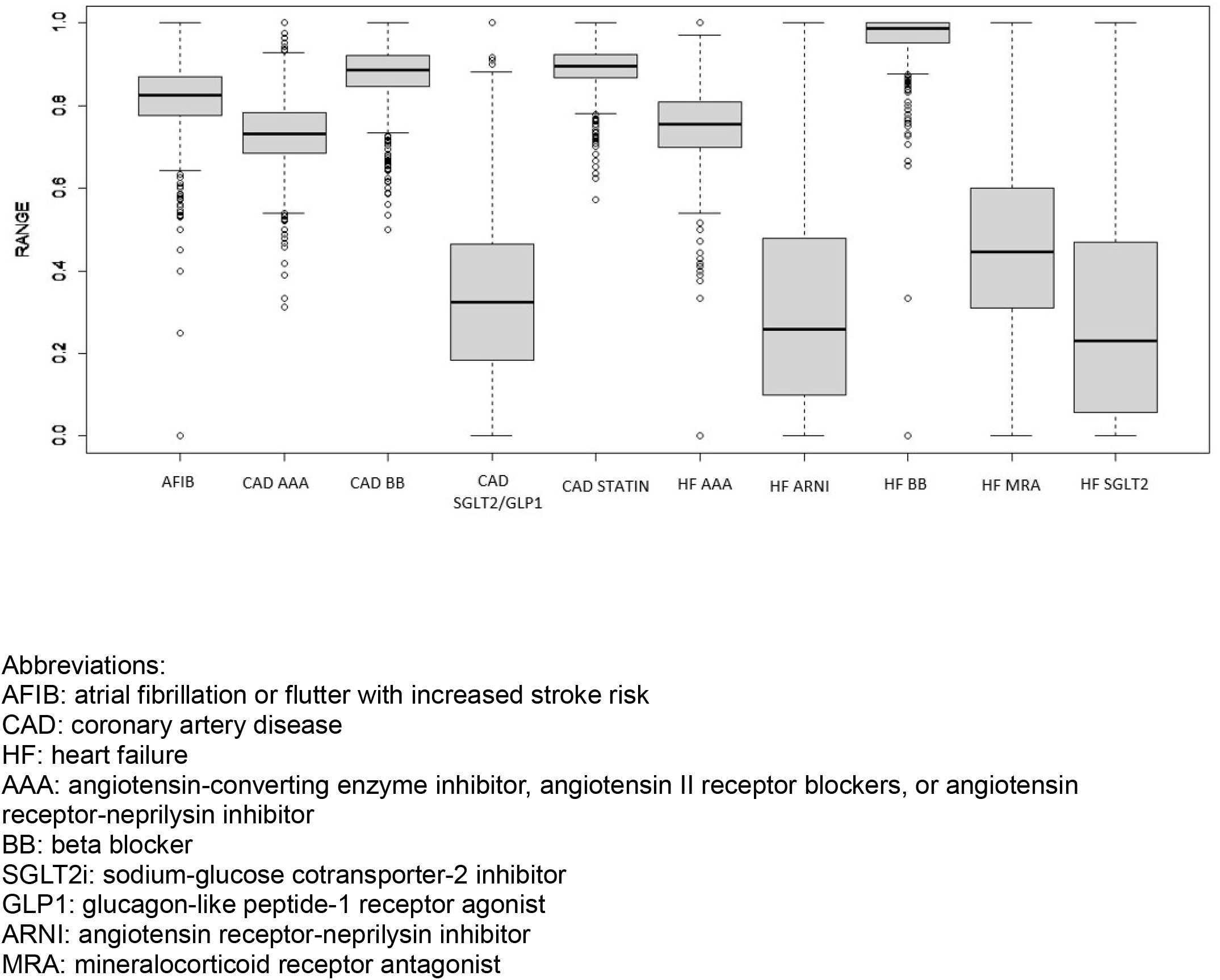
Distribution of clinician achievement of performance measures for patients with AF, CAD, and HFrEF.

Reliability of the performance measures assessed at the clinician level ranged from 0.63 (any ACEi/ARB/ARNI for HFrEF) to 0.90 (SGLT2i or GLP1a for CAD with diabetes). Reliability was higher for the measures based on novel medication use (0.90 for SGLT2i/GLP1 in CAD, 0.86 for ARNI in HFrEF, and 0.90 for SGLT2i in HFrEF) and lower for established treatments.

There was poor correlation between most individual performance measures when measured at the clinician level. Exceptions included moderate correlations between measures including novel medication classes (SGLT2i, GLP1a, ARNI) (**Table 3**).

**Table 3.** Pearson correlation coefficients assessing correlations in the clinician-level achievement of performance measures. Higher values indicate greater correlation in measure achievement, maximum 1.0.

| Condition |  | AF | CAD |  |  |  | HFrEF |  |  |  |  |
| --- | --- | --- | --- | --- | --- | --- | --- | --- | --- | --- | --- |
|  |  | AC | CAD | AAA | BB | SGLT2i /GLP | AAA | ARNI | BB | MRA | SGLT2i |
| AF | AC |  |  |  |  |  |  |  |  |  |  |
| CAD | Statin | 0.13 |  |  |  |  |  |  |  |  |  |
|  | AAA | 0.17 | 0.15 |  |  |  |  |  |  |  |  |
|  | BB | 0.16 | 0.14 | 0.31 |  |  |  |  |  |  |  |
|  | SGLT2i/GLP1 | 0.14 | 0.05 | 0.05 | -0.02 |  |  |  |  |  |  |
| HFrEF | AAA | 0.15 | 0.21 | 0.50 | 0.21 | 0.18 |  |  |  |  |  |
|  | ARNI | 0.08 | 0.02 | 0.14 | -0.04 | 0.56 | 0.20 |  |  |  |  |
|  | BB | 0.14 | 0.16 | 0.06 | 0.23 | -0.005 | 0.13 | 0.02 |  |  |  |
|  | MRA | 0.06 | 0.10 | 0.17 | 0.03 | 0.29 | 0.14 | 0.31 | -0.01 |  |  |
|  | SGLT2i | 0.14 | 0.09 | 0.04 | 0.003 | 0.77 | 0.16 | 0.63 | 0.03 | 0.36 |  |

Abbreviations:
AF: atrial fibrillation or flutter with increased stroke risk
CAD: coronary artery disease
HFrEF: heart failure with reduced ejection fraction
AC: anticoagulation
AAA: angiotensin-converting enzyme inhibitor, angiotensin II receptor blockers, or angiotensin receptor-neprilysin inhibitor
BB: beta blocker
SGLT2i: sodium-glucose cotransporter-2 inhibitor
GLP1: glucagon-like peptide-1 receptor agonist
ARNI: angiotensin receptor-neprilysin inhibitor
MRA: mineralocorticoid receptor antagonist

Median clinician-level achievement of the 10-item composite measure was 72.2%, while the median achievement was 74.6% and 61.5% for similar composites derived from the CAD and HFrEF measures respectively (**Table 1**). Reliability of these composite measures was 0.60, 0.76, and 0.59 respectively.

Case complexity (as estimated by CAN score) had only a modest effect on achievement of performance measures, with an artificial increase of one standard deviation resulting in modest changes in estimated performance ranging from -1.6% (ACEi/ARB/ARNI for HFrEF) to 0.5% (Beta blocker for CAD) (**Table 4**). Similar findings were noted when using Elixhauser Comorbidity Index as an alternative indicator of complexity (Supplemental Appendix).

**Table 4.** Results of a Bayesian experiment assessing the change in performance measure achievement associated with a 1 standard deviation increase in patient complexity, summarized by the VA Care Assessment Needs (CAN) score. Values indicate the estimated change in performance with a one standard deviation increase in CAN score (higher complexity), with negative values indicating worse performance.

| Category | Measure | Direct Causal Effect of Increased Patient Complexity | 89% Confidence Interval |
| --- | --- | --- | --- |
| **AF** | Anticoagulation | 0.07% | 0.02%, 0.12% |
| **CAD** | Statin | -0.64% | -0.99%, -0.35% |
|  | ACEI/ARB/ARNI | -0.32% | -0.52%, -0.15% |
|  | Beta Blocker | 0.53% | 0.19%, 1.02% |
|  | SGT2i/GLP1 | 0.24% | 0.09%, 0.38% |
| **HFrEF** | ACEI/ARB/ARNI | -1.61% | -2.3%, -0.89% |
|  | ARNI | -0.27% | -0.58%, -0.07% |
|  | Beta Blocker | -0.23% | -0.51%, -0.91% |
|  | MRA | -0.13% | -0.22%, -0.01% |
|  | SGLT2i | -0.06% | -0.11%, -0.01% |

Abbreviations:
AF: atrial fibrillation or flutter with increased stroke risk
CAD: coronary artery disease
HFrEF: heart failure with reduced ejection fraction
ACEI: angiotensin-converting enzyme inhibitor
ARB: angiotensin II receptor blockers
ARNI: angiotensin receptor-neprilysin inhibitor
SGLT2i: sodium-glucose cotransporter-2 inhibitor
GLP1: glucagon-like peptide-1 receptor agonist
MRA: mineralocorticoid receptor antagonist

Compared with clinician peer review as a gold standard, 3 representative performance measures demonstrated good sensitivity (0.73-0.95) but poor specificity (0.29-0.50), resulting in balanced accuracy that ranged from no better than chance (0.51 for ACEi/ARB/ARNI for HFrEF) to moderate (0.61 for AF anticoagulation and 0.73 for CAD statin). The CAD and HFrEF measures demonstrated a statistically significant negative directional bias such that the EHR-derived measures were more likely to label an encounter as “not met” relative to clinician peer review (**Table 5**).

**Table 5.** Comparison of EHR-derived performance measure data with OPPE peer review.

|  | Number of peer reviews | Sensitivity | Specificity | Balanced accuracy | Cohen's kappa | Directional bias |
| --- | --- | --- | --- | --- | --- | --- |
| AC for AF | 423 | 0.87<br>(0.82, 0.91) | 0.36<br>(0.21, 0.49) | 0.61<br>(0.54, 0.68) | 0.21<br>(0.07, 0.33) | (-34, 10) |
| Statin for CAD | 647 | 0.95<br>(0.93, 0.97) | 0.50<br>(0.26, 0.75) | 0.73<br>(0.61, 0.85) | 0.29<br>(0.13, 0.44) | (-34, -7) |
| AAA for HFrEF | 521 | 0.73<br>(0.67, 0.77) | 0.29<br>(0.13, 0.48) | 0.51<br>(0.42, 0.60) | 0.004<br>(-0.053, 0.065) | (-142, -88) |

## Discussion

Despite widespread use of performance measures in outpatient cardiology practice, there is remarkably little evidence indicating if and how these measures should be used to assess individual clinician performance. This study addresses multiple knowledge gaps relating to clinician-level measures, assessing measure feasibility, reliability, accuracy, and risk adjustment across multiple metrics for 3 common cardiology conditions. Aligned with National Quality Forum guidelines and using a unique national dataset including over 1,000 clinicians and 700,000 patient encounters, this study identifies strengths and weaknesses of 10 EHR-derived, medication-based performance measures. Overall, our results support the use of clinician-level measures to identify differences in practice among cardiology clinicians and select targets for quality improvement, though they are likely insufficient as standalone tools for clinician quality or competency assessment.

### Feasibility

Reporting burden is a major barrier to the effective use of performance measures. Most quality systems require some form of chart abstraction or manual data entry, even when reporting as electronic clinical quality measures. For example, more than 200 primary care practices across two states were asked to enter EHR data through an online portal to generate electronic quality measures.^24^ The median time to reporting was 7.8 to 10.5 months, with lower-resourced facilities such as Federally Qualified Health Centers reporting slower than hospital-owned practices.

Compared with this process, or the existing case-based peer review system in VA, our EHR-derived measures offer improved timeliness and efficiency. We demonstrated the ability to assess encounter-level achievement for thousands of cardiology visits, assign that achievement to the appropriate cardiology clinician, and aggregate performance over multiple years. Notably, VA has a national EHR with integrated pharmacy data, creating an ideal environment for audit and feedback of medication-based measures. However, integrated pharmacy data is increasingly available in non-VA practices, creating an opportunity to disseminate these strategies beyond VA. Importantly, EHR-based quality measures can improve patient care. A systematic review of EHR quality measures and feedback systems demonstrated feasibility and a positive effect on outcomes in 12 of 14 studies^25^.

### Reliability

Many performance measures have been proposed for the assessment of outpatient cardiovascular practice, and some are actively in use for evaluating physicians participating in CMS pay-for-performance programs or VA practice^10^. These measures are typically based on best practices codified in society guidelines but may not have established evidence of reliability or validity when applied to individual clinicians. The National Quality Forum has recommended testing candidate measures for reliability and validity at the clinician level^20,26^. In the United States there are few health systems that can provide data from a large number of cardiovascular clinicians for appropriate testing of individual-level candidate performance measures. Our national VA cohort includes over 1,000 clinicians including physicians, NPs, and PAs.

Our analysis identified a range of reliability among the 10 candidate measures. Though there is no consensus threshold, some have recommended that reliability exceed 0.8 for comparisons between clinicians, and greater than 0.7 for comparisons between a clinician and the median performance^12,27,28^. We found high reliability for emerging measures based on novel medication classes, such as SGLT2 inhibitors, GLP1 agonists, and ARNIs. As previously documented, uptake of these medications has been heterogeneous, providing an opportunity to identify clinicians who consistently do and do not use these medications^17^. In contrast, several of our 10 performance measures showed relatively low reliability, especially well-established performance measures such as the use of statins for CAD and anticoagulation for AF. These measures had high overall achievement, potentially indicating a ceiling effect and limited ability to discriminate between individual clinicians. Although these measures are important clinical targets, they may be poorly suited for high-stakes assessments of cardiology clinicians due to low reliability.

Furthermore, correlation between different measures at the clinician level was generally poor. Our attempts to develop weighted composite measures based on factor analysis and principle components analysis were not successful^22,23,29^. The opportunity composite measure we ultimately calculated had only modest reliability. These findings challenge the paradigm that clinicians are consistently high or low quality. Instead, our results indicate that cardiology clinicians in VA have individual strengths and weaknesses regarding the use of evidence-based medications.

### Accuracy

In addition to reliability, high-stakes performance measures that impact a clinician’s credentialling, reputation or institutional reimbursement require a high level of validity evidence^30^. One method of assessing validity is by comparison to established peer review processes. In VA, OPPEs are assessed via peer review of a small number of randomly chosen clinical encounters, with reviewers rating the achievement of 3 performance measures (anticoagulation for patients with AF and elevated stroke risk, statin therapy for patients with CAD, ACEI/ARB/ARNI for patients with HFrEF). We replicated this process by recruiting VA clinicians to perform peer review on a subset of our clinical encounters.

In comparison with peer-review, our EHR-derived measures demonstrated balanced accuracy no better than chance for ACEi/ARB/ARNI for HF and only moderate balanced accuracy for AF anticoagulation and statin therapy for CAD. Poor accuracy may indicate that the EHR-derived measures are misclassifying care, or that peer review is not an appropriate “gold standard.” Previous studies have similarly found differences in how claims-based and chart reviewed data classify quality of care. Two small studies of claims-based measures of preoperative cardiac stress testing and carotid revascularization demonstrated that procedures considered “low-value” were sometimes misclassified after chart review was performed.^31,32^

Notably, peer review was only performed for three of the candidate performance measures, which were the established VA measures found to have low reliability and high overall achievement. The low reliability and accuracy of these three performance measures indicate they are poorly suited for high-stakes assessments. It is unknown if the other 7 measures, including those based on novel medication classes, may have better accuracy.

### Risk adjustment

Cardiology practice in VA is heterogeneous, with case-mix differences driven by referral networks, resource availability, and clinician training; sub-specialists, for example, many care for more complex, higher-risk patients who face greater barriers to meeting standard performance measures. It is therefore notable that a Bayesian experiment using our cohort did not find a significant impact of high patient risk (as assessed by established measures of care complexity) on performance measure achievement. While this cannot exclude potential value of risk-adjusting clinician-level measures, it is likely that any adjustment model would provide only modest impact on the classification of individual clinicians based on differences in case mix.

Cardiology practice in VA is heterogenous, with differences in case mix driven by referral networks, resource availability, and clinician training. It is reasonable to speculate that sub- specialists, for example, may care for complex and high-risk patients with greater challenges to achieve standard performance measures. It is therefore notable that a Bayesian experiment using our cohort did not find a significant impact of high patient risk (as assessed by established measures of care complexity) on performance measure achievement. While this cannot exclude potential value of risk-adjusting clinician-level measures, it is likely that any adjustment model would provide only modest impact on the classification of individual clinicians based on differences in case mix.

### Limitations

Our study has several important limitations. First, our measures are based on electronic health data and cannot exclude all patients with contraindications or patient-centered reasons for non-use. However, our methodology included common contraindications to medication use such as allergies, renal insufficiency, and hypotension. Next, we only tested medication-based measures. Other types of measures have been recommended by clinical societies including outcomes (mortality, hospitalization), appropriate management of risk factors (hypertension, lipid levels), and care coordination. However, these measures are more difficult to assess via EHR data and assign to an individual clinician. Our methodology ensured a longitudinal treatment relationship between each patient and clinician dyad and assessed medication prescription measures under the direct control of the clinicians.

### Conclusions

We tested 10 medication-based measures of clinician outpatient care quality for CAD, HFrEF, and atrial fibrillation that can be efficiently measured using EHR data. Several measures had adequate reliability to support comparisons between clinicians, but caution is advised in the use of these measures for high-stakes performance assessment. When compared with peer review, accuracy was only poor to moderate. Additional work is needed to develop reliable and acceptable methods to assess and improve clinician performance. Ultimately, these medication-based performance measures may be useful for clinician benchmarking and the identification of potential care gaps in the context of a comprehensive quality improvement program.

## Acknowledgments

None.

### Sources of Funding

The research was funded by a Veterans Affairs Health Systems Research Merit Award HX003653 (Principal Investigator Dr. Doll).

Dr. Rodriguez was funded by grants from the NIH National Heart, Lung, and Blood Institute (R01HL168188; R01HL167974; R01HL169345) and the Doris Duke Foundation (Grant #2022051).

Dr. Hess reported receiving grants HX002621 from the Veterans Affairs Health Systems Research Career Development Award, HX003570 from the Veterans Affairs Health Systems Research Merit Award outside the submitted work.

### Disclosures

Dr. Rodriguez reports consulting fees from Novartis, Esperion Therapeutics, Edwards, Arrowhead Pharmaceuticals, HeartFlow, iRhythm, Amgen, and Cleerly Health outside the submitted work.

### Supplemental Material

Supplemental Methods

Supplemental Table

### Non-standard Abbreviations and Acronyms

CAD: coronary artery disease
HFrEF: heart failure with reduced ejection fraction
AF: atrial fibrillation
CMS: Center for Medicare and Medicaid Services
MIPS: Merit-Based Incentive Program
VA: Veteran Affairs
EHR: electronic health record
LVEF: left ventricular ejection fraction NP: nurse practitioner
PA: physician assistant
CDW: Corporate Data Warehouse
OPPE: Ongoing Professional Practice Evaluation
SGLT2: sodium glucose cotransporter 2
GLP1: glucagon-like peptide 1 agonists
ARNI: angiotensin receptor-neprilysin inhibitors
CAN: Care Assessment Needs
ACEI: Angiotensin-converting enzyme inhibitors
ARB: angiotensin II receptor blockers
GLP1a: glucagon-like peptide-1 agonist
MRA: mineralocorticoid receptor antagonist

## Data Availability

Following Veterans Affairs data security requirements, data will be shared following a written request to the authors and appropriate approvals from VA.

## Notes

### Competing Interest Statement

The authors have declared no competing interest.

### Author Declarations

We followed the STROKE checklist for a cohort study. The data was de-identified prior to use in this study.

